# Improving sleep after stroke: a randomised controlled trial of digital cognitive behavioural therapy for insomnia

**DOI:** 10.1101/2023.02.07.23285580

**Authors:** Melanie K Fleming, Tom Smejka, Ellie Macey, Ramon Luengo-Fernandez, Alasdair L Henry, Barbara Robinson, Simon D Kyle, Colin A Espie, Heidi Johansen-Berg

**Affiliations:** Wellcome Centre for Integrative Neuroimaging, FMRIB, Nuffield Department of Clinical Neurosciences, University of Oxford, Oxford; Health Economics Research Centre, Nuffield Department of Population Health, University of Oxford, Oxford; Big Health Ltd, London; Sir Jules Thorn Sleep & Circadian Neuroscience Institute, Nuffield Department of Clinical Neurosciences, University of Oxford, Oxford

**Author notes:** Corresponding author: Melanie Fleming, Wellcome Centre for Integrative Neuroimaging, John Radcliffe Hospital, Oxford OX3 9DU, United Kingdom, **E:**. **Contributorship** MKF, TS, HJB designed and conceptualised the study, with input from RLF, ALH, CAE, SDK. TS, EM collected data. MKF, RLF analysed data. MKF and BR drafted the manuscript. All authors reviewed and edited the manuscript and approved the final version.

**Keywords:** CBT, recovery, mood, actigraphy, brain injury, cost-effectiveness

## Abstract

Stroke is frequently accompanied by long-term sleep disruption. We therefore aimed to assess the efficacy of digital cognitive behavioural therapy (dCBT) for insomnia to improve sleep after stroke. A parallel group randomised controlled trial was conducted remotely in participant’s homes/online. Randomisation was online with minimisation of between-group differences in age and baseline Sleep Condition Indicator (SCI-8) score. In total, 86 community dwelling stroke survivors consented, of whom 84 completed baseline assessments (39 female, mean 5.5 years post-stroke, mean 59 years of age) and were randomised to dCBT or control (sleep hygiene information). Follow-up was completed at post-intervention (mean 75 days after baseline) and 8-weeks later. The primary outcome was self-reported insomnia symptoms, as per the SCI-8 (range 0-32, lower numbers indicate more severe insomnia, reliable change 7-points) at post-intervention. There were significant improvements in SCI-8 for dCBT compared to control (intention-to-treat, dCBT n=48, control n=36, 5 imputed datasets, effect of group p≤0.02, η_p_^2^=0.07–0.12, pooled mean difference=-3.35). Additionally, secondary outcomes showed shorter self-reported sleep onset latencies and better mood for the dCBT group, but no significant differences for self-efficacy, quality of life, or actigraphy-derived sleep parameters. Cost-effectiveness analysis found that dCBT dominates over control (non-significant cost savings and higher quality adjusted life years). No related serious adverse events were reported to the researchers. Overall, dCBT for insomnia effectively improves sleep after stroke. Future research is needed to assess earlier stages post-stroke, with a longer follow-up period to determine whether it should be included as part of routine post-stroke care.

Clinicaltrials.gov NCT04272892

## Introduction

Stroke, a leading cause of disability worldwide (Johnson et al., 2019), is frequently accompanied by sleep disruption which persists long-term throughout recovery (Fleming et al., 2021). Insomnia, a sleep disorder characterised by difficulties initiating and maintaining sleep, is highly prevalent in this patient group (pooled prevalence estimate 32%, range 20-70% (Baylan et al., 2020) but studies seeking to improve symptoms are scarce. As poor sleep in the stroke population is associated with depression, fatigue, and reduced quality of life (Byun et al., 2019; Fleming et al., 2021; Tang et al., 2015), this presents an important area for therapeutic consideration. Additionally, sleep difficulties after acquired brain injury, including stroke, have also been correlated with poorer recovery outcomes (Fleming 2021), further highlighting the significance for the need of good sleep in this patient group.

Cognitive behavioural therapy (CBT) is the first-line recommended treatment for insomnia which has been shown to be effective in improving sleep across a range of patient groups, and preliminary efficacy for in-person treatment has been demonstrated following stroke (Herron et al., 2018; Nguyen et al., 2017). However, provision of in-person CBT is limited by scarcity of trained therapists, long-wait lists, and is costly, culminating in high unmet demands for treatment (Koffel et al., 2018). Digital CBT (dCBT) mitigates these limitations and provides an option for delivering treatment at scale. Indeed, Sleepio (an automated dCBT programme) is effective at improving insomnia (Espie et al., 2012), mood (Luik et al., 2017) and cognitive function (Kyle et al., 2020). Although the effects of stroke, such as reduced mobility or pain, may generate potential barriers to completing standard dCBT techniques, we have previously demonstrated that with some additional information supplied, Sleepio can be feasibly used by community dwelling stroke survivors (Smejka et al., 2022). However, many stroke survivors also experience long term difficulties with movement, language, and cognition, which are factors that could potentially impact the effectiveness of a behavioural intervention. As such, it is important to determine the efficacy of dCBT in this population specifically before recommending its use.

This study aimed to assess the efficacy of dCBT to improve sleep in chronic stroke survivors. We hypothesised that dCBT would result in greater improvements to sleep than provision of sleep hygiene information, and that these improvements would be sustained at least 8-weeks later. Our secondary aims were to assess the effects on mood, quality of life and self-efficacy, as well as actigraphy-derived sleep parameters. Finally, to evaluate the real-world impact of improved sleep for this population, we explored the cost-effectiveness of dCBT.

## Methods

### Design

This was a two-arm parallel group, randomised controlled trial comparing dCBT with provision of sleep hygiene information. The study was approved by the University of Oxford Central University Research Ethics Committee (R40803) and registered as a clinical trial prior to enrolment of the first participant (*clinicaltrials.gov* NCT04272892).

### Participants

To be eligible, participants had to be: 1) aged >18 years; 2) > 3months post-stroke; 3) interested in improving their sleep; 4) living in the UK with reliable internet access; 5) able to understand verbal and written English (with assistance from carer if needed); and 6) able and willing to provide informed consent. Participants were excluded if they: 1) had a serious clinical condition that could affect participation in the study, including scheduled surgery in the next 5 months; 2) were currently undergoing a psychological treatment programme for insomnia; 3) were pregnant; 4) had uncontrolled seizures (contraindication to sleep restriction included as part of dCBT); 5) had untreated diagnosed obstructive sleep apnoea; or 6) did habitual shift-work.

Participants were recruited from the community between February 2020 and June 2021, by advertising through UK stroke and brain injury charities, stroke user/support groups, social media, and our research database. After receiving the information sheet and having the opportunity to discuss the study with a researcher, participants provided written informed consent online (using the Jisc platform). All participants (regardless of group allocation) received online shopping vouchers as compensation for their time (£15 per assessment time-point, max £45).

### Outcomes

Outcome assessments were collected at baseline, at the end of treatment (herein termed “post-intervention”), and at a follow-up 8-weeks later. Participants completed assessments online, except the EuroQol 5-dimension questionnaire (EQ-5D) and actigraphy which were posted to their home.

The primary outcome was insomnia symptoms post-intervention, which was assessed using the Sleep Condition Indicator (SCI-8, max score 32). The SCI-8 was developed to evaluate insomnia based on the 5^th^ edition of the Diagnostic and Statistical Manual of Mental Disorders (DSM-5), with higher values indicating fewer insomnia symptoms. The initial validation study identified that scores ≤16 indicate probable insomnia (Espie et al., 2014). To our knowledge, the SCI-8 has not been specifically validated in a stroke population, but previous studies have identified significant differences in scores for people with stroke (Fleming et al., 2021) and brain injury (Fleming et al., 2020) in comparison with age- and sex-matched controls.

Secondary sleep outcomes included the SCI-8 at the 8-week follow-up, sleep onset latency (SOL; from the online sleep diary recorded during the first and last week of the intervention period), and actigraphy-derived sleep measures (estimated total sleep time, wake after sleep onset, sleep fragmentation; Supplementary Methods 1.5) at post-intervention and at the 8-week follow-up. The actigraphy device used was a Motionwatch-8 (Camntech Ltd, UK), worn on the participant’s least-affected wrist for 7-nights per time-point. As we opted to include participants whose sleep problems were not severe enough to be considered probable insomnia, which could limit the effect size observed, we also intended to assess SCI-8 score post-intervention for the sub-sample of participants who had probable insomnia (SCI-8 ≤ 16) at baseline (Espie et al., 2014).

Other secondary outcomes included mood, as assessed by the Patient Health Questionnaire (PHQ-9, max score 27; Kroenke et al., 2001), and the Generalized Anxiety Disorder questionnaire (GAD-7, max score 21; Spitzer et al., 2006); and self-efficacy as per the stroke self-efficacy questionnaire (SSE, max score 130; Jones et al., 2008). Quality of life was assessed with the EQ-5D-5L™ (Herdman et al., 2011), and the short form stroke impact scale (SF-SIS, max index value 100; Jenkinson et al., 2013).

### Resource use and unit costs

Participants completed a bespoke version of the Client Service Receipt Inventory (Beecham & Knapp, 2001) to categorise National Health Service (NHS) resource use over the 8-weeks prior to randomisation and over the 8-week follow-up period (Supplementary Methods 1.1).

The per-patient cost to the NHS of Sleepio was £45. Unit costs were obtained from the Personal Social Services Research Unit’s publication for 2020 (Curtis & Burns, 2020) and the NHS Schedule of Reference Costs for 2020.

### Randomisation

Following baseline assessment, participants were randomised to intervention (dCBT) or control (sleep hygiene information) using freely available online software (rando.la), with minimisation of factors age and baseline SCI-8 score to attempt to ensure balance across the two groups. MFK held access to the randomisation software, and upon completion of each participant’s baseline assessment, entered their SCI-8 score and age. Group allocation was then recorded onto a spreadsheet for the research assistant to access, and to inform the participant of their allocation. Thus, the study team did not know which treatment would be assigned prior to recruitment.

Due to the nature and practicalities of the intervention, it was not possible to blind participants or the research team to group allocation. However, a blind-to-hypothesis approach was used whereby participants were told that the study was testing sleep improvement interventions, but not which group was anticipated to show greater effects. Those who analysed the data (MKF, RLF) had minimal interactions with participants throughout the intervention period.

### Intervention Group

Participants were given access to Sleepio (www.sleepio.com), comprising of 6 weekly online sessions (each 15-20 minutes) delivering evidence-based CBT techniques for insomnia, and completion of a daily sleep diary for the intervention period (Supplementary Methods 1.2). Based on feedback received during our previous qualitative usability study of Sleepio (Smejka et al., 2022), participants were also emailed a document at the beginning of the intervention period which provided additional information to aid using the programme in the context of stroke. In instances in which participants chose not to complete all CBT sessions, they were asked to complete the post-intervention assessment as soon as possible.

### Control group

Participants were emailed a sleep hygiene brochure containing suggestions on lifestyle and environmental factors associated with sleep disturbance (Supplementary Methods 1.3), and completed a daily online sleep diary for one week at the beginning and end of the intervention period.

### Sample size

The sample size calculation was based on the primary outcome (SCI-8 score). We intended to collect 68 complete datasets (1:1 ratio). Based on the between-group effect size estimate of d=1.2 (Espie et al., 2012) it was initially determined that 24 participants were required (α=0.05, power 80%). However, as we opted to include people with sleep difficulties who would not meet the criteria for clinical insomnia (based on the SCI-8), and anticipated a more modest effect (Smejka et al., 2022) (d=0.7), we thus determined that 68 full datasets were required. Allowing for withdrawal, we aimed to enrol 86 participants. We anticipated this would enable a full group analysis for the primary outcome, and a secondary subgroup analysis including only participants with probable insomnia (as per the SCI-8).

When 56 participants had been recruited, it was clear that withdrawal from the dCBT group was such that maintaining a 1:1 randomisation would lead to insufficient dCBT participants for 68 complete datasets at 1:1 ratio. The remaining 30 participants were thus randomised at 2:1 (treatment:control).

### Analysis

#### Statistics

Data for the primary outcome (SCI-8 score post-intervention) were analysed using intention-to-treat (ITT) with multiple imputation of missing values (Supplementary Methods 1.4). We conducted an analysis of covariance (ANCOVA), with the dependent variable of SCI-8 post-intervention, fixed factor of Group (dCBT, control), and covariates of baseline SCI-8 and sex.

Secondary outcomes were analysed as a complete case ITT population, restricted to randomised participants for whom data was available at all time-points. This was chosen to limit analysis to participants who engaged in the intervention to some extent, even if they did not complete the programme. Mixed ANCOVAs were used with the within-subject factor of time (post-intervention and 8-week follow-up), between-subject factor of group, and baseline score as a covariate. When group effects were found, chi square tests were used to explore differences in the proportion of participants reaching the reliable or minimally clinically important change (MCID). Two-sided p-values (significance p<0.05) are reported with estimated effect sizes (partial eta-squared: η_p_^2^) where appropriate.

#### Cost-effectiveness

EQ-5D-5L responses were converted into utility values (van Hout et al., 2012). As no deaths were reported, individual quality adjusted life years (QALYs) were estimated by combining utility estimates. For the cost-effectiveness analysis, differential mean QALYs were adjusted for baseline utility, sex and age using ordinary least squares regression.

Costs were compared using a t-test. For the cost-effectiveness analysis, differential mean costs were adjusted for baseline costs, age, and sex. To evaluate if dCBT was cost-effective, an incremental analysis was carried out, with the mean cost difference between groups divided by the mean QALY difference to give the incremental cost-effectiveness ratio (ICER). The main analysis used a healthcare perspective, but sensitivity analyses also included informal care costs. As per NICE recommendations, we judged an intervention to be cost-effective if the ICER was ≤£20,000 per QALY gained.

The non-parametric percentile method was used for calculating the confidence interval around the ICER, using 10,000 bootstrap estimates of the mean cost and QALY differences. The cost-effectiveness acceptability curve was used to show the probability that dCBT is cost-effective at a threshold of £20,000 per QALY gained, and for different values of the willingness to pay for an additional QALY.

#### Exploratory mediation analysis

To further understand the effects of dCBT for insomnia on mood and actigraphy-derived sleep disruption measures, we explored whether differences in these secondary outcomes were mediated by SCI-8 score changes using the “mediation” package in R (Supplementary Methods 1.6).

## Results

### Demographics

Recruitment took place between February 2020 and June 2021, with a total of 86 participants consented as planned. The trial ended in December 2021, once all follow-up assessments were completed. Two participants withdrew without completing the baseline (no reason given). The remaining 84 participants (mean (SD) age 58.6 (13.5) years, 39 female, mean (SD) 5.5 (5) years post-stroke) were randomised (Table 1). Based on the SCI-8, 83% reported having sleep problems for >6 months.

**Table 1.** Baseline demographics and assessment values

|  | dCBT | Control |
| --- | --- | --- |
| N | 48 | 36 |
| Age, years | 58.5 (12.7) | 58.7 (14.7) |
| Sex (female): N (%) | 17 (35.4%) | 22 (61.1%) |
| Years since most recent stroke | 6 (5) | 6 (5) |
| Number of strokes, N (%) |  |  |
| 1 | 41 (85%) | 30 (83%) |
| > 1 | 7 (15%) | 6 (17%) |
| **Relevant Medications** |  |  |
| Antidepressant |  |  |
| *SSRI* | 8 (17%) | 3 (8%) |
| *SNRI* | 2 (4%) | 2 (6%) |
| *SARI* | 1 (2%) | 0 (%) |
| *TCA* | 2 (4%) | 3 (8%) |
| *Other* | 1 (2%) | 1 (3%) |
| Z-drug | 0 (0%) | 3 (8%) |
| Gabapentinoid | 8 (17%) | 5 (14%) |
| Melatonin Receptor Agonist | 1 (2%) | 1 (3%) |
| **Patient reported outcomes** |  |  |
| N | 48 | 36 |
| SCI-8 | 12.0 (6.3) | 11.4 (7.0) |
| *Probable Insomnia*: N (%) | 38 (79.2%) | 30 (83.3%) |
| PHQ-9 | 10.2 (4.4) | 9.5 (5.1) |
| GAD-7 | 8.9 (5.5) | 7.6 (5.4) |
| SF-SIS | 47.3 (13.2) | 43.1 (13.7) |
| SSE | 85.8 (28.6) | 73.5 (36.5) |
| **Actigraphy** |  |  |
| N | 48 | 34 |
| Estimated total sleep time<br>(hours:min) | 7:01 (1:08) | 7:26 (1:22) |
| Wake After Sleep Onset (min) | 57 (28) | 63 (34) |
| Sleep fragmentation index | 31.5 (13.8) | 32.1 (15.7) |

Values are Mean (Standard Deviation) unless otherwise specified. SSRI= selective serotonin reuptake inhibitor, SNRI= Serotonin and norepinephrine reuptake inhibitor, SARI= Serotonin antagonist and reuptake inhibitor, TCA=tricyclic antidepressant, Z-drug e.g. Zopiclone or Zolpidem

Sixteen participants (13 dCBT, 3 control) withdrew without completing the post-intervention assessment (Figure 1; Supplementary Table S1). Comparison of characteristics between participants who withdrew and those who completed the post-intervention assessment showed that there was no difference for baseline SCI score (|t|(82)=0.16, p = 0.874) or age (|t|(82)=0.17, p = 0.869). However, participants who withdrew had significantly worse symptoms of depression (PHQ-9; |t|(82)=2.46, p = 0.016) and anxiety (GAD-7; |t|(82)=2.25, p = 0.027) at baseline than those who completed the study, and lower self-efficacy (SSE; |t|(82)=2.84, p = 0.006).

**Figure 1:**
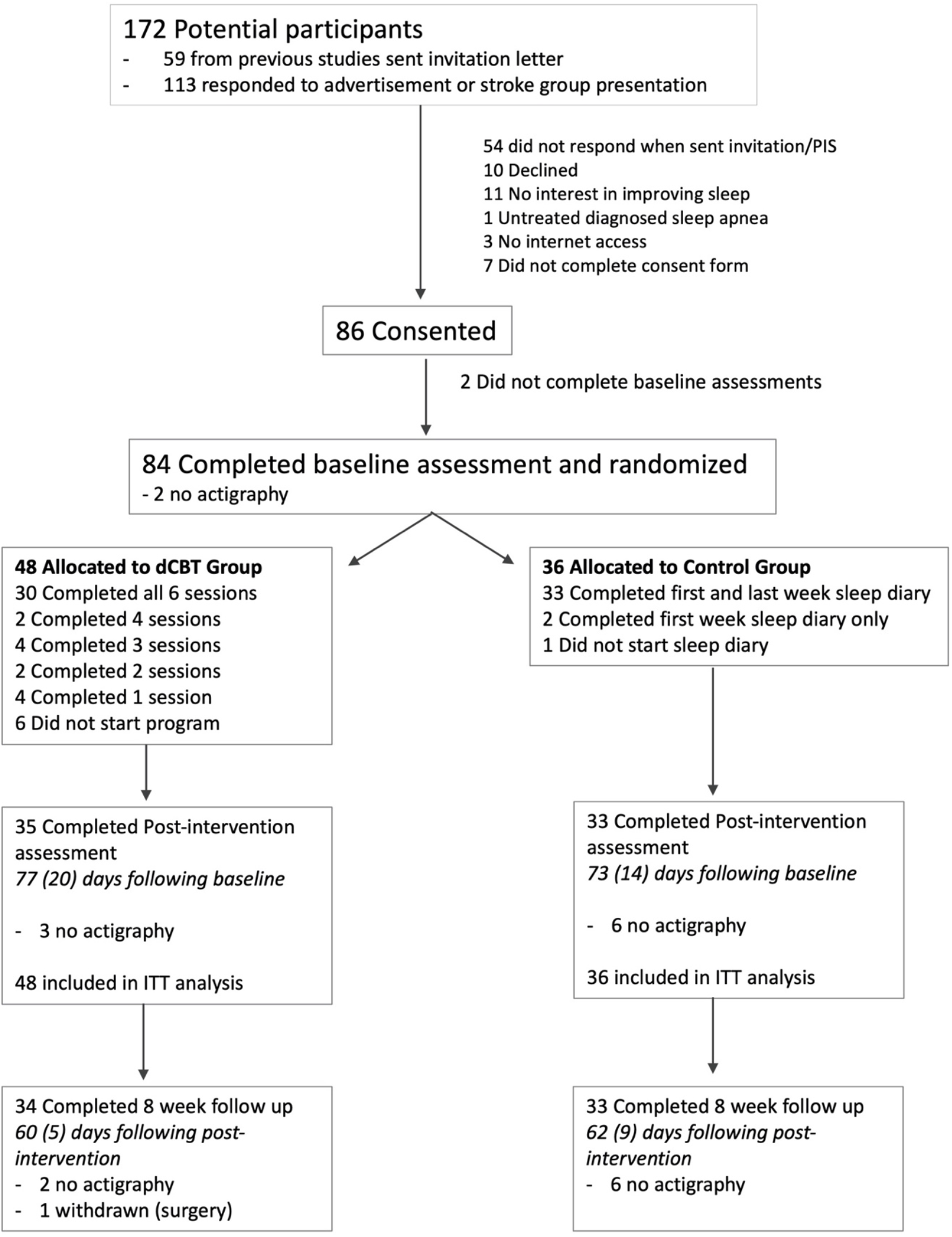
Consort diagram. For the primary outcome (SCI-8 score post-intervention), intention-to-treat (ITT) analysis was with imputation of missing data. For secondary outcomes, complete case analysis was used. PIS=participant information sheet. dCBT=digital cognitive behavioural therapy for insomnia. Mean (standard deviation) days between assessments did not differ between groups.

We estimated it would take 6-8 weeks for participants to complete the dCBT programme and attempted to match control group assessment timing on an ongoing basis. Although the time from baseline to post-intervention was on average longer than anticipated (Figure 1), timeframes did not differ between groups (p>0.2).

### Sleep Condition Indicator

The primary outcome, SCI-8 score post-intervention (ITT dCBT n=48, control n=36) was significantly greater following dCBT than control, adjusted for baseline SCI-8 and sex, across original and imputed datasets (Table 2; Supplementary Table S2), with a medium effect size. This is indicative of fewer symptoms of insomnia following dCBT.

**Table 2.** Intention-to-treat analysis of primary outcome (SCI-8 post-intervention).

| | Control | | dCBT | | Adjusted Mean<br>Difference*<br>(95% CI) | Effect of Group | Effect<br>size<br>($\eta_p^2$) |
| --- | --- | --- | --- | --- | --- | --- | --- |
|  | Baseline | Post-intervention | Baseline | Post-intervention |  |  |  |
| Original data | 11.4 (7.0) | 14.9 (7.3) | 12.0 (6.3) | 18.5 (5.9) | -3.661 (-6.14, -1.18) | $F_{1,64}=8.72$,<br>$p=0.004$ | 0.12 |
| Imputation 1 | - | 14.7 (7.0) | - | 18.2 (6.0) | -3.40 (-5.64, -1.16) | $F_{1,80}=9.09$,<br>$p=0.003$ | 0.10 |
| Imputation 2 | - | 14.6 (7.1) | - | 18.5 (6.7) | -3.65 (-5.91, -1.38) | $F_{1,80}=10.27$,<br>$p=0.002$ | 0.11 |
| Imputation 3 | - | 14.8 (7.2) | - | 17.6 (6.7) | -2.77 (-5.09, -0.46) | $F_{1,80}=5.67$,<br>$p=0.020$ | 0.07 |
| Imputation 4 | - | 14.5 (7.1) | - | 18.3 (5.7) | -3.65 (-5.92, -1.37) | $F_{1,80}=10.19$,<br>$p=0.002$ | 0.11 |
| Imputation 5 | - | 14.6 (7.2) | - | 18.1 (6.1) | -3.26 (-5.48, -1.05) | $F_{1,80}=8.60$,<br>$p=0.004$ | 0.10 |
| **Pooled** |  | **14.6** |  | **18.1** | **-3.35** | - | - |

Our secondary objective in relation to the SCI-8 was to test for maintenance of effects. There was an effect of group (Mixed ANCOVA: F_1,64_=6.35, p=0.014, η_p_^2^=0.09, medium effect), and no group by time interaction (F_1,64_=0.74 p=0.39), suggesting improved SCI-8 score for dCBT across post-intervention and the 8-week follow-up timepoints (Figure 2A).

**Figure 2.**
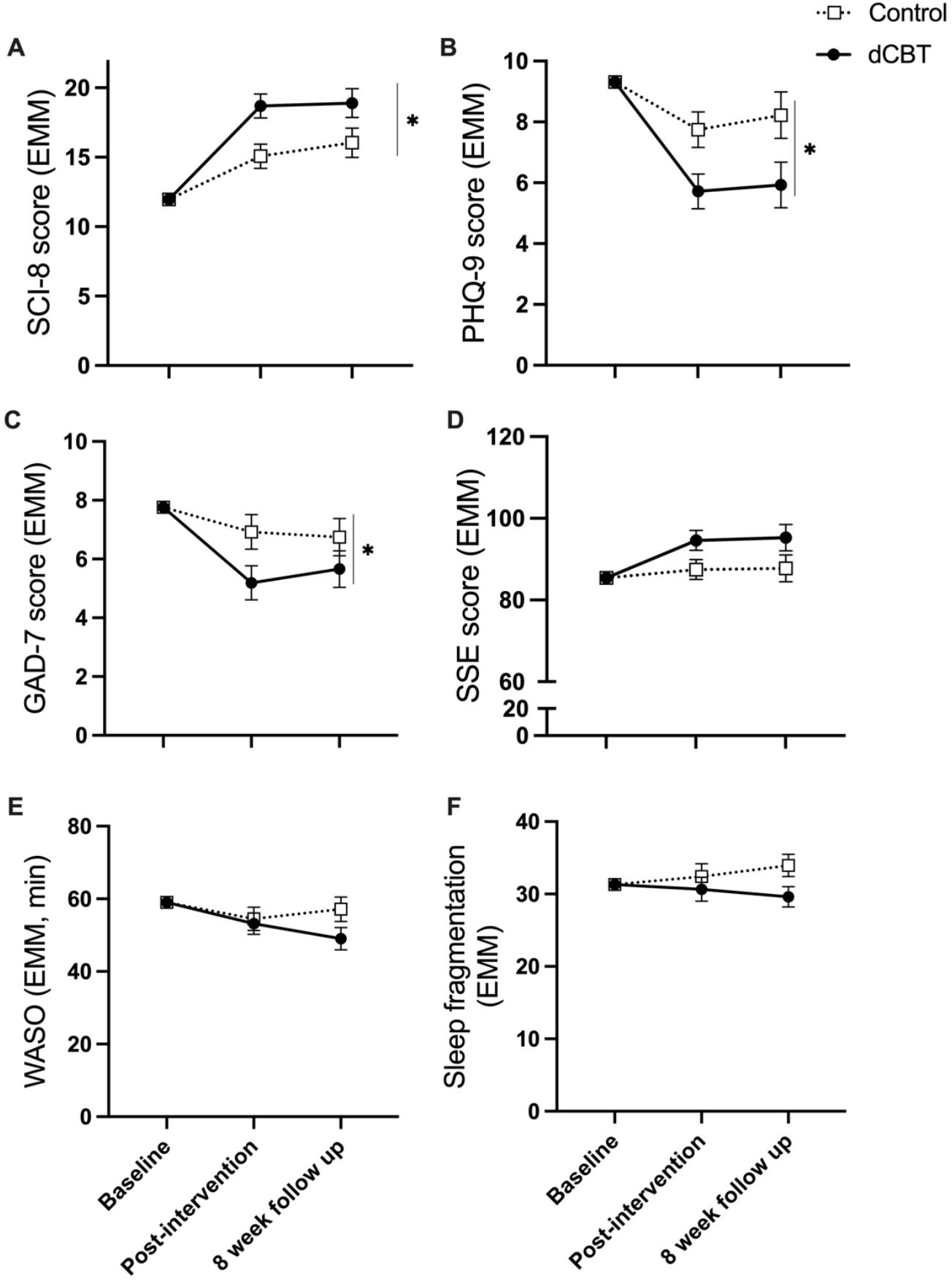
Estimated marginal means (baseline score covaried) at post-intervention and 8-week follow-up for questionnaire and actigraphy measures, **A:** Sleep Condition Indicator (higher values indicate fewer symptoms of insomnia), **B:** Patient Health Questionnaire (lower scores indicate fewer symptoms of depression), **C:** Generalised Anxiety Disorder questionnaire (lower scores indicate fewer symptoms of anxiety), **D:** Stroke Self-Efficacy scale (higher scores indicate better self-efficacy), **E**: Wake after sleep onset (higher time is more wakefulness during sleep period). **F**: Sleep fragmentation index (higher values indicate more disrupted sleep). Error bars are standard error of the mean. * significant effect of group (adjusted means were better for dCBT than control, p<0.05).

For the secondary sub-group analyses including only participants with probable insomnia, based on the SCI-8 score at baseline (SCI-8 ≤16; dCBT n=29, control n=27), there was an effect of group (F_1,52_=8.27, p=0.006, η_p_^2^=0.14, medium effect), as adjusted SCI-8 was higher for dCBT than control. As an additional, post-hoc analysis, we also found that a larger proportion of the dCBT group (baseline ≤16) scored >16 at post-intervention, suggesting symptom resolution (71% *vs* 30%, χ^2^(1)=9.61, p=0.002). For visualisation purposes, the proportions of participants reaching the criteria for probable insomnia at each timepoint are in Supplementary Figure S1.

### Secondary patient reported outcomes

Group means can be found in Supplementary Tables S3-S7. For depression (PHQ-9) and anxiety (GAD-7) there were significant group effects (PHQ-9: F_1,64_=6.754, p=0.012, η_p_^2^=0.095, medium effect; GAD-7: F_1,64_=4.109, p=0.047, η_p_^2^=0.060, medium effect), and no group by time interactions (PHQ-9: F_1,64_=0.079, p=0.780; GAD-7: F_1,64_=0.405, p=0.527), suggesting improved mood following dCBT compared to control across follow-up timepoints (Figure 2B,C).

For SOL, taken from the online sleep diary, the ANCOVA (with first week as covariate) showed a significantly shorter sleep latency for dCBT than control at the end of the intervention period (F_1,63_=6.406, p=0.014, η_p_^2^=0.092, medium effect).

For self-efficacy (SSE) there was a non-significant effect of group (F_1,64_=3.990, p=0.050, η_p_^2^=0.059, medium effect; Figure 2D), and no group by time interaction (F_1,64_=0.009, p=0.923).

For quality of life, there was no effect of group for SF-SIS (F_1,64_=0.132, p=0.718), nor group by time interaction (F_1,64_=0.827, p=0.367). Similarly, there were no between-group differences for EQ-5D utilities or visual analogue scale (VAS) scores (see Supplementary Table S5 for adjusted mean differences; p>0.05).

### Costs and cost-effectiveness

Over the 8-week follow-up period, and after including the costs of Sleepio, the dCBT group had non-significantly lower NHS care costs than control (adjusted mean difference -£349, 95% CI: -1,035 to 337, p=0.31, Supplementary Table S8). After inclusion of informal care costs, the adjusted mean cost difference was -£330 (95% CI: -1,550 to 891, p=0.59).

From a health-care perspective, dCBT was dominant over sleep hygiene information (i.e. overall cost savings and associated with higher QALYs – Table 3). The probability that dCBT was cost-saving was 0.876. The probability increased to 0.885 and 0.911 at £20,000 and £100,000 per QALY gained threshold respectively.

**Table 3.**
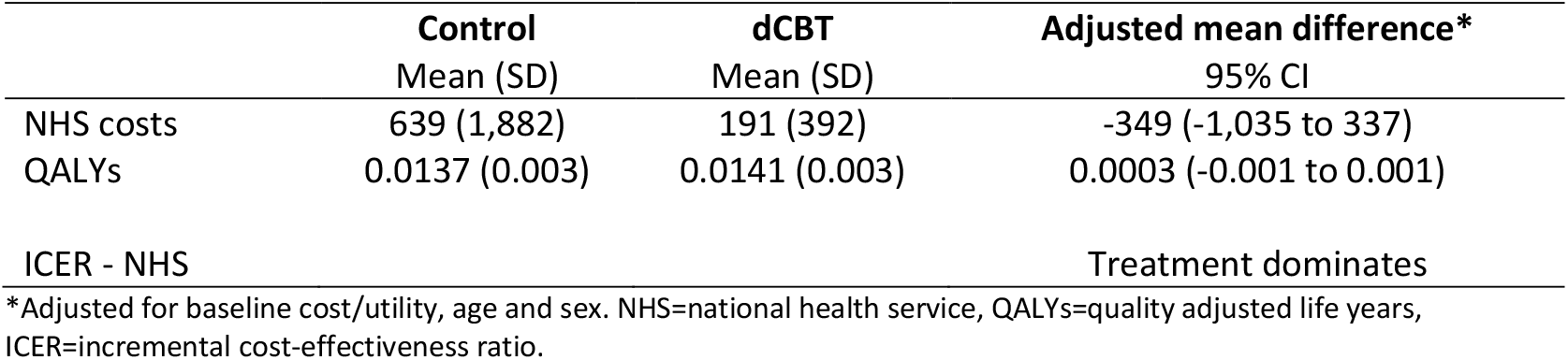
Cost-effectiveness of dCBT (treatment) compared to control.

### Secondary actigraphy outcomes

There were no group effects (all F_1,54_<2.6, p>0.1), nor group by time interactions (all F_1,54_<2.8, p>0.09) for any of the analysed actigraphy parameters (Figure 2E, F; Supplementary Table S3).

### Estimate of importance of difference

For outcomes where a significant group effect was observed, we assessed the proportion of participants meeting the reliable or minimally clinically important difference (MCID). There is no established MCID for SCI-8, but more dCBT participants experienced a reliable change at post-intervention (≥7 points (Espie et al., 2018), 49% *vs* 21%, χ^2^(1)=5.57, p=0.018) which was not statistically significant at the 8-week follow-up (50% *vs* 30%, χ^2^(1)=2.70, p=0.100).

Similarly, significantly more dCBT participants reached the MCID for PHQ-9 and GAD-7 at post-intervention (PHQ-9: ≥5 points (Kroenke et al., 2001), 40% *vs* 15%, χ^2^(1)=5.21, p=0.022; GAD-7: ≥4 points (Toussaint et al., 2020), 43% *vs* 12%, χ^2^(1)=7.97, p=0.005) but this was not significant at the 8-week follow-up (PHQ-9: 32% *vs* 15%, χ^2^(1)=2.73, p=0.099; GAD-7: 35% *vs* 15%, χ^2^(1)=3.588, p=0.058).

### Exploratory mediation analyses

The effect of group on PHQ-9 at post-intervention was mediated via the SCI-8 score (Figure 3A). The average causal mediation effect (ACME) was -1.18 [95% CI: -2.25 to -0.37], p=0.002. This suggests that reductions in depression are mediated by improvements in insomnia symptoms. No mediation effects were observed for GAD-7 (ACME -0.33 [95% CI: -1.06 to 0.30], p=0.256).

**Figure 3.**
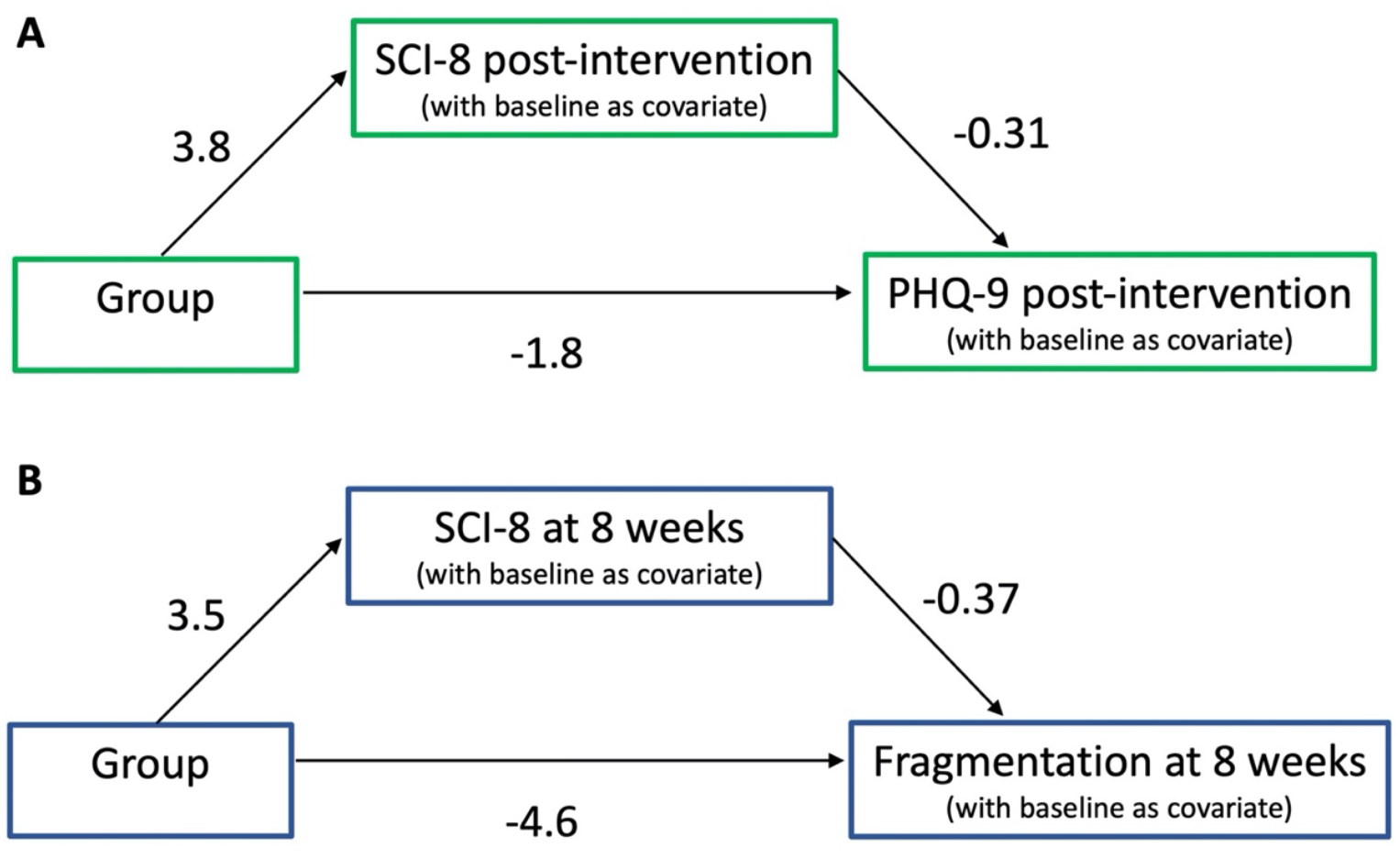
Exploratory mediation analyses. **A:** The SCI-8 score mediates the effect of group on PHQ-9. The Average Causal Mediation Effect (ACME) was -1.18 (n=68). **B:** The ACME for sleep fragmentation was -1.24 (n=59).

Finally, given the (non-significant) visual tendency for lower wake after sleep onset (WASO) and sleep fragmentation for dCBT at the 8-week follow-up (see Figure 2E,F), we explored whether SCI-8 mediated apparent changes in sleep disruption from actigraphy. The ACME was -1.47 [95% CI: -5.04 to 1.26], p=0.264 for WASO and -1.24 [95% CI: -3.26 to 0.01], p=0.054 for sleep fragmentation.

### Adverse Events

No related serious adverse events were reported to the research team. Other adverse effects related to study participation are in the Supplementary Results 2.5.

## Discussion

This study found significantly fewer symptoms of insomnia for community dwelling stroke survivors following dCBT for insomnia compared to provision of sleep hygiene information. Digital CBT for insomnia, specifically Sleepio, is effective across a range of clinical populations, and this is the first study to demonstrate efficacy after stroke. Results of the cost-effectiveness analyses also suggest that dCBT has the potential to be cost-effective in this population.

Insomnia symptoms are highly prevalent after stroke (Baylan et al., 2020), and poor sleep is also a risk factor for stroke (Gottlieb et al., 2019). Here we demonstrate improvements in SCI-8 score following dCBT, with an overall medium effect size across ITT and complete case analyses. Participants seeking to improve sleep were included, regardless of severity, to increase generalisability. This may have limited the magnitude of our effect, but previous research indicates some efficacy of dCBT in treating sub-threshold insomnia symptoms (Denis et al., 2020). Our sub-analysis restricted to participants with probable insomnia (based on baseline SCI-8 score) demonstrated similar findings to the full group analysis. We found a reliable change for approximately half of the dCBT group who completed the follow-up assessments. This is broadly comparable with Ymer et al (2021), who found clinically meaningful improvements in sleep quality for people with acquired brain injury (60% of participants post-intervention, 46% at 2-month follow-up). There is growing evidence that improvements conferred by CBT are sustained for at least 6-12 months (Luik et al., 2020; van der Zweerde et al., 2019). Nevertheless, given the long-term nature of stroke, future studies with a longer follow-up are needed to ascertain whether repeat or refresher sessions will be required to prevent return of sleep problems. Additionally, as we deliberately included community dwelling, rather than hospitalised, stroke survivors, it is not anticipated that the findings translate directly to the early stages of stroke recovery. Further studies are needed to evaluate and implement evidence-based treatments for sleep in acute and rehabilitation stroke units.

Mood disorders are highly prevalent after stroke (Jørgensen et al., 2016; Rafsten et al., 2018), and we found fewer symptoms of depression and anxiety following dCBT for insomnia. Our exploratory mediation analysis suggests that improvements in depression are mediated to some extent by improvements in SCI-8, as seen in adults without stroke (Henry et al., 2021). Sleep may therefore serve as a treatment target to improve mental health in this population. However, it is important to consider the inherent relationships between self-reported measures, e.g. individuals low in mood or confidence may rate their sleep as worse. This is particularly relevant given that there were no significant actigraphy changes.

A recent meta-analysis demonstrated that there are typically minimal or no improvements in actigraphy or polysomnography outcomes following CBT (Mitchell et al., 2019). Although reasons for this are unclear, it may be partly explained by studies recruiting via self-reported rather than objective measures, as insomnia is defined by self-reported complaints. For the current study, actigraphy data was not available from all participants (reasons in Supplementary Results 2.6), and although no statistical significance was found there is some apparent visual tendency towards improved sleep fragmentation index and WASO at 8-weeks following dCBT (Figure 2). It may be that improvements in actigraphy parameters could develop after initial changes in sleep habits and perception in this patient group. However, this is entirely speculative and requires adequately powered studies with a longer follow-up to investigate. Nonetheless, our exploratory analyses are encouraging, suggestive of a tendency towards less sleep fragmentation with improvements in SCI-8.

### Limitations

Despite conducting a qualitative study to understand and address usability concerns (Smejka et al., 2022), a substantial proportion of participants still withdrew. The dropout was comparable to that found previously in people without stroke (Espie et al., 2012; Ho et al., 2015; Seyffert et al., 2016). Withdrawn participants exhibited lower mood and self-efficacy at baseline, which is consistent with studies from other populations using in-person and digital CBT (Ong et al., 2008; Yeung et al., 2015). Since active engagement is required, first addressing feelings of low mood and confidence may be beneficial. A hybrid model, whereby dCBT is combined with clinician input, may help participants to discuss options and receive support for self-managing treatment. Indeed, hybrid treatment was effective in a sample of 52 people with stroke or traumatic brain injury (Ford et al., 2022).

We used ITT analysis for the primary outcome but chose to use complete case analyses for the secondary measures. We acknowledge that this introduces bias but felt it important to examine changes in secondary outcomes specifically for stroke survivors who were able and willing to engage in the intervention to some extent. The current study therefore provides an initial assessment of outcomes which may or may not be responsive to dCBT in this population, but researchers in the future should use these results to guide study design to attempt replication.

As we did not have access to brain imaging or medical records it is unknown whether stroke lesion characteristics impacted on sleep or treatment response, or whether stroke characteristics differed between groups. The effect observed here may also be limited by undiagnosed comorbid sleep disorders, e.g. sleep apnoea, although CBT for insomnia remains effective in people with comorbid obstructive sleep apnoea (Sweetman et al., 2017). We were unable to complete in-person sleep assessments and relied on participant report for diagnoses of sleep apnoea. Though we acknowledge these are clear limitations, we nevertheless demonstrate significant sleep improvements in this cohort.

Results of the cost-effectiveness analyses should be interpreted cautiously given the relatively small sample and short follow-up duration. Nevertheless, the promising results are consistent with previous studies demonstrating cost-effectiveness (Darden et al., 2021), and should be extended in a larger trial.

## Conclusion

Cognitive behavioural therapy is the first line recommendation for treatment of insomnia and here we provide evidence of efficacy of dCBT in community-dwelling stroke survivors. More research is needed to ascertain who is most likely to benefit, the extent to which efficacy is similar earlier after discharge from hospital, and how long effects persist.

## Supporting information

CONSORT checklist

## Data Availability

All data produced are available online at https://osf.io/zmvfx/

## Acknowledgements

Thank you to the stroke groups and charities who disseminated advertisements and invited us to join group meetings. Thanks to Piergiorgio Salvan and Marin Beims for assistance with manuscript preparation.

## Data availability

The data that support the findings of this study are openly available on the Open Science Framework at https://osf.io/zmvfx/.

## Rights retention

This research was funded in whole, or in part, by the Wellcome Trust [Grant number 110027/Z/15/Z]. For the purpose of open access, the author has applied a CCBY public copyright license to any Author Accepted Manuscript version arising from this submission.

