## Supplementary for "Improving sleep after stroke: a randomised controlled trial of digital cognitive behavioural therapy for insomnia"

### 1 Supplementary Methods

*1.1 Resource use*

We used a bespoke version of the Client Service Receipt Inventory to record resource use. At baseline, participants retrospectively recalled resource use over the preceding 8-weeks. After the post-intervention assessment, participants were provided a logbook to record resource use and any changes in medications over the time until the 8-week follow-up assessment.

We included the following resource use categories over the 8-week periods: hospitalizations; outpatient consultations with psychiatrists or other hospital consultants; day hospital visits; primary care visits; visits with therapists; social worker visits; and day care contacts. Outside the health and social care perspective adopted, we also asked patients to complete the nature and amount of any informal care (i.e. unpaid care from relatives, friends or neighbours) received.  Unit costs for consultations in primary care, social worker visits and day care contacts were obtained from the Personal Social Services Research Unit's publication for 2020 (Curtis & Burns, 2020). For all other contacts, unit costs were derived from the NHS Schedule of Reference Costs for 2020 (*NHS Digital. Reference Costs*, 2020).

*1.2 Sleepio programme*

Cognitive behavioral therapy (CBT) sessions within Sleepio include cognitive (cognitive restructuring, positive imagery, paradoxical intention, putting the day to rest) and behavioural interventions (stimulus control therapy, sleep restriction therapy, relaxation), and an educational component (sleep hygiene, psycho-education). Individuals are instructed to keep a daily sleep diary (accessed in the programme). This includes questions about the time they went to bed and got up, how long it took them to fall asleep (sleep onset latency), number and duration of awakenings and a rating of the quality of their sleep. The sleep diary, in addition to additional questions throughout the programme, aids personalisation of treatment content. They have access to additional content through the Sleepio "Library".

Participants were also emailed a document at the beginning of the intervention period to provide additional information to aid using the programme, based on feedback received during our previous qualitative usability study with stroke survivors (Smejka et al., 2022). This document included screenshots to demonstrate how to find the sleep diary and the CBT sessions and some possible modifications to behavioural advice given. Specifically, this included 1) Napping: If you are unable to stop napping completely, we would suggest trying to have a short nap (10-15 minutes) around 2-3pm but no later. 2) Sleep-bed connection: If moving to another room is difficult for you, we would suggest sitting up on the edge of your bed (on the covers if you can) instead of leaving the room, then lying back down when you feel sleepy again. 3) Progressive muscle relaxation: If you experience painful muscle tension on the affected side of your body, or find it difficult to relax again after tensing a particular muscle (or group of muscles), then we would suggest only using parts of your body that you can do this comfortably (e.g the side of your body that wasn't affected by the stroke).

*1.3 Control group information*

Participants were emailed a sleep hygiene brochure at the beginning of the intervention period. The sleep hygiene brochure was created based on suggestions freely available on the internet, in particular from the National Sleep Foundation (https://www.thensf.org/sleep-health-topics/). This included suggestions to limit daytime napping (to 30 minutes), increase exercise and natural light during the day, avoid stimulants, create a pleasant sleeping environment and a regular routine. Participants also completed an online sleep diary (with questions matching those of the dCBT group) for one week at the beginning, and approximately 6 weeks later (matched to the dCBT group timings on an ongoing basis). The control group were given free access to Sleepio following completion of the final follow up assessment, but no data was collected from this for the study.

*1.4 Multiple imputation procedure*

We used multiple imputation in SPSS (V28, IBM inc.) with the predictor variables of SCI-8, SSE, GAD-7 and PHQ-9 at baseline, and imputed values for SCI-8 post-intervention (values constrained 0-32). Sixteen values were missing (3 control, 13 dCBT) and 5 imputed datasets created.

*1.5 Actigraphy analysis*

The Motionwatch 8 contains an accelerometer and motion is converted into an activity count. Data were averaged by the monitor into 30 s epochs. Sleep measures were extracted using the custom software, Motionware (Camntech Ltd). Since it was not always possible to rely on the event marker in this population, the time that participants tried to sleep (e.g. turned off the light) and got up were taken from a short sleep diary (based on questions 2 and 6 from the consensus sleep diary) completed each day whilst wearing the actigraphy monitor, and adjusted based on the apparent movement and time of the event marker if present. The median value of the 7 nights was obtained for sleep measures: estimated total sleep time (minutes), wake after sleep onset (WASO; minutes), and the fragmentation index (time mobile relative to immobile). We chose to focus predominantly on measures of sleep disruption (WASO, fragmentation) as these measures have previously been highlighted to be different for stroke survivors compared to controls (Fleming et al., 2021). We did not assess sleep onset latency or sleep efficiency since it was not always possible to obtain an accurate indication of time in bed prior to the onset of sleep.

*1.6 Exploratory mediation analyses*

We wanted to further understand the effects of dCBT for insomnia on measures of anxiety, depression and actigraphy derived sleep disruption measures. To do this, we ran exploratory mediation analyses using the “lm” and “mediation” packages within R (Tingley et al., 2014). The mediation package combines the linear regression outcome and mediator models and estimates the average causal mediation effect.

For mood measures, since the results showed no group by time interaction, we chose to focus just on the primary timepoint (post-intervention). The outcome model was a linear model with PHQ-9 or GAD-7 as the dependent variable, Group (dCBT, control) as the independent variable, SCI-8 score post-intervention as the mediator, and baseline values as covariates. The mediator model was a linear model with SCI-8 score post-intervention as the dependent variable, Group as the independent variable, and baseline values for SCI-8 and PHQ-9/GAD-7 as covariates.

For actigraphy measures, Figure 2 shows a visual tendency towards a group difference at the 8-week follow-up for WASO and Sleep fragmentation index (although group by time interaction was not significant). We therefore chose to explore whether variability in WASO and fragmentation at the 8-week follow-up was mediated by SCI-8 score at this timepoint. Similar to mood measures, the outcome model was a linear model with WASO or fragmentation index as the dependent variable, Group (dCBT, control) as the independent variable, SCI-8 score at 8-weeks as the mediator, and baseline values as covariates. The mediator model was a linear model with SCI-8 score at 8-weeks as the dependent variable, Group as the independent variable, and baseline values for SCI-8 and WASO/fragmentation as covariates. A quasi-Bayesian approximation was used for confidence intervals, with 1000 simulations.

*1.7 Adverse effects*

The protocol approved by the research ethics committee specified two expected adverse effects:

1. Increased daytime sleepiness as a result of the sleep restriction that is part of the CBT programme. Participants were advised in the participant information sheet that a possible risk of taking part in the study is tiredness initially following reduced sleep as part of an adjusted sleep schedule. They were told that this sleep restriction is part of the Sleepio programme and is thought to aid sleep overall. They were advised to avoid driving or using dangerous machinery if feeling excessively tired, and that should they need to, short daytime napping before approximately 3pm may be of use. It was recommended that they contact the researchers if concerned at any point.
2. Participant answers to questions on the PHQ-9 and the GAD-7 could identify concerns regarding mood (e.g depressive symptoms or anxiety). It is made clear on the participant information sheet that these questionnaires are used for research purposes, not for diagnostic purposes. Anyone who scored 10 points or more on either of these questionnaires was sent an email advising them of contacts, such as the National Health Service and mental health charities, should they wish to talk to someone.

We did not specifically ask participants for information about adverse events at any point in the study.

#### 2. Supplementary Results

*2.1 Participant withdrawal*

Participants were not obliged to provide a reason for withdrawal, and were encouraged to complete the follow-up assessments, even if they did not complete the intervention. Nonetheless, 13 dCBT (2 female) and 3 control (2 female) participants withdrew following randomization. Reasons provided for withdrawal are in Supplementary table S1.

**Supplementary Table S1:** Reasons for withdrawal from study

| Reason provided | Number of participants |
|---|---|
| Sleep problems too mild to complete programme | 3 |
| Started pharmacological treatment for sleep and didn't want to continue | 2 |
| Unwell | 1 |
| Didn't think Sleepio was the right option for them (didn't sign up) | 1 |
| Young children mean they didn't think they could do the programme (didn't sign up) | 1 |
| No reason given | 8 |

One participant withdrew following the post-intervention assessment as they were admitted to hospital for surgery (pre-planned but not scheduled at the time of recruitment to the study).

**Supplementary Table S2:** Group estimated marginal mean and 95% confidence interval for intention-to-treat analysis of primary outcome (SCI-8 post-intervention adjusted for baseline SCI-8 and sex)

| | **Control** *Mean (95% CI)* | **dCBT** *Mean (95% CI)* | **Effect size^** *Cohen's D* |
|---|---|---|---|
| Original data | 14.88 (13.12 to 16.64) | 18.54 (16.83 to 20.25) | 0.7 |
| Imputation 1 | 14.78 (13.11 to 16.45) | 18.18 (16.74 to 19.62) | 0.6 |
| Imputation 2 | 14.72 (13.03 to 16.40) | 18.37 (16.91 to 19.82) | 0.7 |
| Imputation 3 | 14.78 (13.06 to 16.51) | 17.56 (16.07 to 19.04) | 0.5 |
| Imputation 4 | 14.57 (12.88 to 16.27) | 18.22 (16.76 to 19.68) | 0.7 |
| Imputation 5 | 14.73 (13.08 to 16.38) | 17.99 (16.57 to 19.42) | 0.6 |
| **Pooled** | **14.72 (13.05 to 16.39)** | **18.06 (16.47 to 19.66)** | |

^To facilitate future study planning and comparisons with other treatments for sleep disturbance in the future, we computed Cohen's D effect size. To do this, we first performed a linear regression with dependent variable SCI-8 post-intervention and independent variables SCI-8 at baseline and sex, then the unstandardized residuals were entered into independent samples t-tests (one for each imputation) using SPSS.

##### *2.2 Presence of insomnia*

The main results demonstrate clear improvements in SCI-8 after dCBT and 71% of the dCBT group (who had probable insomnia at baseline) scored better than the cut-off score for probable insomnia after the intervention. We therefore wanted to (post-hoc) better understand the proportion of our sample who met the cut-off for probable insomnia over time. Figure S1 demonstrates the relative changes in proportions of participants scoring ≤16 or >16 on the SCI-8 at each time-point, for visualization purposes only.

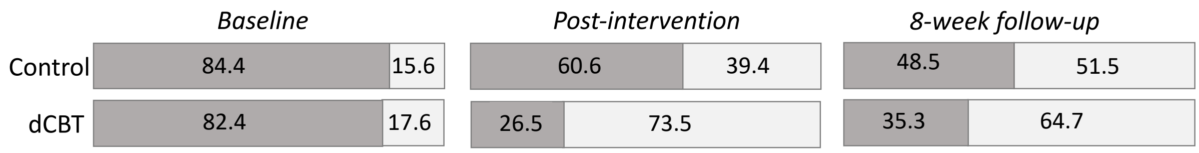


**Supplementary Figure S1.** Percentage of participants in each group scoring ≤16 (probable insomnia; dark grey) or >16 (light grey) at each assessment time-point. Top = control group, bottom = dCBT group. Data are from complete cases; dCBT n=34, control n=33. Note that this was a post-hoc quantification, for exploratory purposes.

**Supplementary Table S3:** Demographics and outcome data for complete cases

| | Baseline | | Post-intervention | | 8-week follow-up | | Mean Difference* |
|---|---|---|---|---|---|---|---|
| | Control | dCBT | Control | dCBT | Control | dCBT | [95% CI] |
| **Demographics** | | | | | | | |
| Age, years | 58.8 (14.5) | 58.2 (11.9) | | | | | |
| Years since stroke | 5 (5) | 6 (6) | | | | | |
| Sex (female; N (%)) | 20 (60.6%) | 15 (42.9%) | | | | | |
| **Patient reported outcomes** | | | | | | | |
| SCI-8 | 11.7 (7.0) | 12.2 (5.4) | 14.9 (7.3) | 18.9 (5.6) | 15.8 (8.3) | 19.1 (7.3) | -3.24 [-5.81 to -0.67] |
| PHQ-9 | 8.8 (4.7) | 9.8 (4.3) | 7.4 (5.4) | 6.1 (4.5) | 7.8 (6.6) | 6.4 (5.4) | 2.16 [0.50 to 3.82] |
| GAD-7 | 7.0 (5.1) | 8.5 (5.2) | 6.3 (5.6) | 5.8 (4.6) | 6.2 (5.3) | 6.2 (5.2) | 1.41 [0.02 to 2.80] |
| SF-SIS | 43.6 (14.0) | 50.4 (13.2) | 47.2 (15.1) | 55.1 (15.3) | 48.9 (15.0) | 55.1 (14.9) | -0.65 [-1.21 to 2.92] |
| SSE | 77.2 (34.8) | 93.4 (22.9) | 80.4 (33.6) | 101.4 (22.9) | 81.3 (34.6) | 101.5 (23.6) | -7.33 [-14.66 to 0.00] |
| SOL^ | 37.6 (39.5) | 41.5 (67.6) | 27.4 (25.7) | 19.1 (27.6) | - | - | 9.84 [2.07 to 17.61] |
| **Actigraphy** | | | | | | | |
| Estimated total sleep time (h:min) | 7:25 (1:22) | 7:21 (0:54) | 7:23 (1:16) | 7:01 (0:44) | 7:10 (1:20) | 7:09 (0:57) | 0:09 [-0:11 to 0:28] |
| Fragmentation index | 31 (17) | 31 (14) | 32 (17) | 31 (16) | 34 (19) | 30 (15) | 3.03 [-0.80 to 6.85] |
| Wake after sleep onset (min) | 55 (22) | 63 (31) | 51 (21) | 56 (30) | 54 (26) | 52 (30) | 4.7 [-3.0 to 12.5] |

Values are Mean (Standard deviation) unless specified. Questionnaires: dCBT n=34, control n=33, except SOL dCBT n=34, control n=32. Actigraphy: dCBT n=31, control n=26. *There was no group by time interaction, value is therefore the mean group difference adjusted for baseline (control - dCBT). ^ sleep onset latency is the median value over the first and last week of the intervention period. Adherence to the sleep diary was good, 90% of participants had an available sleep diary for at least 6 nights at each time-point.

2.3 *Sex differences*

There is an increased awareness of the need of research to be aware of and try to further our understanding of differences in treatment responses between men and women (Rexrode et al., 2022). For information purposes, and to facilitate future study planning, Table S4 therefore presents the group means (complete cases) for the primary measure (SCI-8 score post-intervention, adjusted for baseline) separated by sex.

**Supplementary Table S4.** Group means for SCI-8 post-intervention, adjusted for baseline, separated by sex.

| | **Male** | | | **Female** | | |
|---|---|---|---|---|---|---|
| | **Control** | **dCBT** | **Adjusted* mean difference** | **Control** | **dCBT** | **Adjusted* mean difference** |
| **n** | 13 | 20 | | 20 | 15 | |
| **SCI-8** | 14.8 (12.5 to 17.1) | 18.7 (16.8 to 20.5) | -3.9 (-1.0 to -6.8) | 14.9 (12.2 to 17.6) | 18.4 (15.2 to 21.5) | -3.5 (-0.7 to -7.6) |

Values are mean with 95% confidence interval. * mean group difference (control - dCBT) with baseline covaried. Data were available from 33 control and 35 dCBT participants at the post-intervention assessment.

*2.4 PHQ-9 scoring*

The PHQ-9 includes a question relating to sleep ("Over the last 2 weeks how often have you been bothered by… trouble falling/staying asleep, sleeping too much"). If this question was removed from scoring, then the effect of group remained ($F_{1,64}$=6.01, p=0.017, $\eta^2$=0.086), and there remained no interaction between group and time ($F_{1,64}$=0.439, p=0.510).

**Supplementary Table S5**. EQ-5D utility and VAS scores at each time-point by treatment group

| | **Control** mean (SD) | **dCBT** mean (SD) | **Difference** mean (95% CI) | **Adjusted Difference*** mean (95% CI) |
|---|---|---|---|---|
| **Baseline** | | | | |
| EQ-5D utility | 0.55 (0.27) | 0.59 (0.22) | 0.04 (-0.07 to 0.15) | |
| EQ-5D score | 63 (20) | 58 (24) | -5 (-15 to 5) | |
| **Post-intervention** | | | | |
| EQ-5D utility | 0.63 (0.16) | 0.65 (0.19) | 0.02 (-0.06 to 0.11) | 0.02 (-0.05 to 0.10) |
| EQ-5D score | 65 (18) | 68 (19) | 2 (-8 to 11) | 4 (-3 to 11) |
| **8-week follow up** | | | | |
| EQ-5D utility | 0.61 (0.22) | 0.65 (0.17) | 0.03 (-0.06 to -0.14) | 0.02 (-0.05 to 0.09) |
| EQ-5D score | 67 (20) | 69 (17) | 2 (-7 to 12) | 6 (-1 to 12) |

*adjusted for baseline utility/score, age and sex.

**Supplementary Table S6.** Responses to the EQ-5D-5L questionnaire at baseline, post-intervention and 8-week follow-up in the control group

| | **Baseline** n (%) | **Post-intervention** n (%) | **8-week follow-up** n (%) |
|---|---|---|---|
| **Mobility** | | | |
| No problems | 5 (14) | 6 (20) | 5 (16) |
| Slight problems | 11 (31) | 11 (37) | 10 (31) |
| Moderate problems | 14 (39) | 11 (37) | 14 (44) |
| Severe problems | 5 (14) | 2 (7) | 2 (6) |
| Unable to walk | 1 (3) | 0 | 1 (3) |
| **Self-care** | | | |
| No problems | 10 (28) | 12 (40) | 12 (38) |
| Slight problems | 15 (42) | 13 (43) | 12 (38) |
| Moderate problems | 6 (17) | 3 (10) | 6 (19) |
| Severe problems | 3 (8) | 2 (7) | 2 (6) |
| Unable to self-care | 2 (6) | 0 | 0 |
| **Usual activities** | | | |
| No problems | 4 (11) | 6 (20) | 5 (16) |
| Slight problems | 12 (33) | 12 (40) | 14 (44) |
| Moderate problems | 13 (36) | 9 (30) | 9 (28) |
| Severe problems | 4 (11) | 3 (10) | 4 (13) |
| Unable to undertake | 3 (8) | 0 | 0 |
| **Pain/discomfort** | | | |
| No pain/discomfort | 8 (22) | 5 (17) | 7 (22) |
| Slight pain/discomfort | 16 (44) | 18 (60) | 17 (53) |
| Moderate pain/discomfort | 9 (25) | 5 (17) | 6 (19) |
| Severe pain/discomfort | 2 (6) | 2 (7) | 1 (3) |
| Extreme pain/discomfort | 1 (3) | 0 | 1 (3) |
| **Anxiety/depression** | | | |
| Not anxious/depressed | 13 (36) | 13 (43) | 18 (56) |
| Slightly anxious/depressed | 16 (44) | 13 (43) | 8 (25) |
| Moderately anxious/depressed | 4 (11) | 3 (10) | 5 (16) |
| Severely anxious/depressed | 2 (6) | 1 (3) | 1 (3) |
| Extremely anxious/depressed | 1 (3) | 0 | 0 |

**Supplementary Table S7.** Responses to the EQ-5D-5L questionnaire at baseline, post-intervention and 8-week follow-up in the dCBT group

| | **Baseline**<br>n (%) | **End of treatment**<br>n (%) | **8-week follow-up**<br>n (%) |
|---|---|---|---|
| **Mobility** | | | |
| No problems | 15 (31) | 11 (34) | 9 (29) |
| Slight problems | 7 (15) | 8 (25) | 9 (29) |
| Moderate problems | 19 (40) | 7 (22) | 9 (29) |
| Severe problems | 7 (15) | 6 (19) | 4 (13) |
| Unable to walk | 0 | 0 | 0 |
| **Self-care** | | | |
| No problems | 21 (44) | 19 (59) | 14 (45) |
| Slight problems | 16 (33) | 8 (25) | 13 (42) |
| Moderate problems | 8 (17) | 4 (13) | 3 (10) |
| Severe problems | 1 (2) | 1 (3) | 1 (3) |
| Unable to self-care | 2 (4) | 0 | 0 |
| **Usual activities** | | | |
| No problems | 7 (15) | 8 (25) | 9 (29) |
| Slight problems | 15 (31) | 9 (25) | 8 (26) |
| Moderate problems | 15 (31) | 9 (28) | 9 (29) |
| Severe problems | 9 (19) | 5 (16) | 5 (16) |
| Unable to undertake | 2 (4) | 1 (3) | 0 |
| **Pain/discomfort** | | | |
| No pain/discomfort | 12 (25) | 10 (31) | 9 (29) |
| Slight pain/discomfort | 19 (40) | 13 (41) | 13 (42) |
| Moderate pain/discomfort | 13 (27) | 7 (22) | 7 (23) |
| Severe pain/discomfort | 3 (6) | 2 (6) | 2 (6) |
| Extreme pain/discomfort | 1 (2) | 0 | 0 |
| **Anxiety/depression** | | | |
| Not anxious/depressed | 13 (27) | 13 (41) | 12 (39) |
| Slightly anxious/depressed | 21 (44) | 11 (34) | 12 (39) |
| Moderately anxious/depressed | 11 (23) | 7 (22) | 6 (19) |
| Severely anxious/depressed | 3 (6) | 1 (3) | 1 (3) |
| Extremely anxious/depressed | 0 | 0 | 0 |

**Supplementary Table S8**. Care costs (£) in the 8-weeks before randomisation and following the intervention

| | **Control** mean (SD) | **dCBT** mean (SD) | **Difference** mean (95% CI) | **Adjusted difference*** mean (95% CI) |
|---|---|---|---|---|
| **Baseline (8 weeks before randomisation)** | | | | |
| Hospitalisations | 708 (3000) | 153 (1057) | -555 (-1484 to 373) | |
| Visits to: | | | | |
| Psychiatrist | 0 | 0 | 0 | |
| Other consultant | 33 (100) | 49 (106) | 16 (-29 to 62) | |
| GP | 34 (64) | 32 (45) | -2 (-26 to 22) | |
| Psychiatric nurse | 3 (12) | 0 | -3 (-6 to 1) | |
| Therapist | 43 (108) | 53 (147) | 10 (-48 to 68) | |
| Social worker | 1 (9) | 0 | -1 (-4 to 1) | |
| Day hospital | 23 (135) | 135 (454) | 113 (-43 to 269) | |
| Day care | 3 (17) | 0 | -3 (-8 to 2) | |
| Total cost NHS | 845 (3,261) | 419 (1,197) | -426 (-1,442 to 559) | |
| Informal care | 1,831 (3,574) | 2,182 (5,323) | 350 (-1,692 to 2,393) | |
| TOTAL COST | 2,677 (5,054) | 2,600 (5,349) | -76 (-2,368 to 2,216) | |
| **8 week follow up period** | | | | |
| Hospitalisations | 304 (1,233) | 58 (237) | -246 (-690 to 197) | -220 (-676 to 234) |
| Visits to: | | | | |
| Psychiatrist | 75 (431) | 0 | -75 (-227 to 77) | -71 (-221 to 78) |
| Other consultant | 111 (306) | 37 (83) | -74 (-186 to 38) | -76 (-169 to 17) |
| GP | 39 (57) | 32 (47) | -7 (-34 to 20) | -5 (-34 to 25) |
| Psychiatric nurse | 8 (36) | 0 | -8 (-20 to 5) | -8 (-20 to 5) |
| Therapist | 30 (79) | 17 (74) | -14 (-52 to 24) | -12 (-47 to 23) |
| Social worker | 0 | 0 | 0 | 0 |
| Day hospital | 74 (237) | 51 (199) | -23 (-132 to 85) | -17 (132 to 97) |
| Day care | 0 | 0 | 0 | 0 |
| Sleepio | 0 | 45 | 45 (N/A) | 45 (N/A) |
| Total cost NHS | 639 (1,882) | 191 (392) | -448 (-1,126 to 231) | -349 (-1,035 to 337) |
| Informal care | 1,435 (3,072) | 1,619 (4,707) | 183 (-1,780 to 2,148) | -120 (-1,425 to 1,185) |
| TOTAL COST | 2,074 (3,538) | 1,810 (4,723) | -264 (-2,328 to 1,800) | -330 (-1,550 to 891) |

*adjusted for baseline costs, age and sex. SD=standard deviation, CI=confidence interval, GP=general practitioner

*2.5 Related and/or expected adverse effects*

A high proportion of the participants (n=50) scored ≥10 on the PHQ-9 or GAD-7 questionnaires, suggestive of clinically significant symptoms of depression and/or anxiety at one or more assessments. As planned in the protocol, they were therefore sent a letter recommending that they contact their General Practitioner if they would like to talk to someone, as well as providing links to mental health charities which they could contact for support.

One participant (control group) reported that wearing the actigraphy monitor and recording the sleep diary made them anxious and they felt they slept worse; as such we recommended that they discontinue the actigraphy assessment. Additionally, two participants reported experiencing slight skin irritation from the actigraphy monitor.

One participant (dCBT group) reported that the sleep restriction week was mentally and physically draining, that they were very tired and unable to work during the week. They reported feeling it had lasting damaging effects on their mind and body for two weeks. Nevertheless, they did complete the study and their SCI-8 score improved by 1-point at post-intervention and by a further 5-points at the 8-week follow-up.

One participant (dCBT group) had a substantial worsening of their SCI-8 score (12-point decrease at post-intervention, which improved by 3-points at the 8 week-follow up but remained lower than baseline). They had reported to the researchers during the intervention that they were unwilling to follow all aspects of the programme and were unwilling to complete the programme (but remained in the study to complete the questionnaire assessments).

*2.6 Reasons for Unavailable Actigraphy Data*

Actigraphy data was not available from all participants. The reasons for this include: not worn due to slight skin irritation (n=2), technical issues (n=2), lost monitor (n=2).

3. **Supplementary References**


Curtis, L., & Burns, A. (2020). *Unit Costs of Health and Social Care 2020, .* https://www.pssru.ac.uk/project-pages/unit-costs/unit-costs-2020/

Fleming, M. K., Smejka, T., Henderson Slater, D., Chiu, E. G., Demeyere, N., & Johansen-Berg, H. (2021). Self-Reported and Objective Sleep Measures in Stroke Survivors With Incomplete Motor Recovery at the Chronic Stage. *Neurorehabilitation and Neural Repair*, *35*(10), 851–860. https://doi.org/10.1177/15459683211029889

*NHS Digital. Reference costs*. (2020). https://digital.nhs.uk/data-and-information/data-collections-and-data-sets/data-collections/reference-costs

Rexrode, K. M., Madsen, T. E., Yu, A. Y. X., Carcel, C., Lichtman, J. H., & Miller, E. C. (2022). The Impact of Sex and Gender on Stroke. *Circulation Research*, *130*(4). https://doi.org/10.1161/CIRCRESAHA.121.319915

Smejka, T., Henry, A. L., Wheatley, C., Espie, C. A., Johansen-Berg, H., & Fleming, M. K. (2022). A qualitative examination of the usability of a digital cognitive behavioral therapy for insomnia program after stroke. *Brain Injury*, 1–8. https://doi.org/10.1080/02699052.2022.2034182

Tingley, D., Yamamoto, T., Hirose, K., Keele, L., & Imai, K. (2014). Mediation: R package for causal mediation analysis. *Journal of Statistical Software*, *59*(5). https://doi.org/10.18637/jss.v059.i05
